# The neuropathology of “dementia epileptica”

**DOI:** 10.64898/2026.04.29.26352020

**Authors:** Silke Emilie Laustsen, Janette Krogh Petersen, Camilla Dyremose Cornwall, Martin Wirenfeldt, Christoph Patrick Beier

## Abstract

**Objective:** Accumulation of hyperphosphorylated tau (pTau) is a proposed mechanism for dementia associated with epileptic seizures but the evidence directly linking seizures and pTau accumulation remains weak. Here, we aimed at determining the burden of pTau pathology in a historical cohort of epilepsy patients who developed dementia hypothesizing an association between seizure onset zone and pTau accumulation.

**Methods:** Post-mortem brain tissue was obtained from the Danish Brain Collection comprising autopsies from psychiatric patients that died between 1945-1982. Sections from the middle frontal gyrus, thalamus, and medial temporal lobe from both hemispheres were stained for pTau and beta-amyloid and quantified by a blinded assessor. Comparisons were conducted using non-parametric tests.

**Results:** Thirty-two patients (median age 61.5 years, 59.4% men) were included. pTau pathology was detected in 22 brains (68.8%), with Tau Burden Scores (0-108) ranging from 2 (almost undetectable) to 94 (high load; median 6.5). pTau burden was significantly associated with age at death (Spearman’s rho = 0.62, *p* < 0.001) and duration of epilepsy (Spearman’s rho = 0.47, *p* = 0.02), but not with other clinical variables. Among 11 patients with focal seizures, a significantly higher pTau (*p* = 0.02) but not a higher beta-amyloid burden (*p* = 1.0) was observed in the epileptogenic hemisphere. 81.3% of all patients (n=26) had a competing dementia diagnosis: three patients fulfilled the pathological criteria of Alzheimer’s dementia, 13 patients had clinical and/or autoptic diagnosis of vascular dementia.

**Significance:** In this cohort, dementia epileptica was associated with increased pTau burden in the epileptogenic hemisphere and time since diagnosis supporting the concept of seizure-induced pTau accumulation. However, pTau is unlikely to be the primary neuropathological link between epilepsy and dementia in the cohort studied given the overall mild pathology and competing diagnoses.

## Introduction

Epilepsy, characterized by recurrent and unprovoked seizures, affects approximately 50 million people worldwide (1, 2). People with epilepsy (PWE) often present with cognitive, psychiatric, and behavioral comorbidities, which may have a greater impact on their health and quality of life than the seizures themselves (3). Cognitive deficits are observed in 70-80% of patients, ranging from subtle difficulties in attention and memory to severe cognitive impairments and dementia (4).

The connection between epilepsy and progressive cognitive decline has been acknowledged since the late 19^th^ century, when it was referred to as “dementia epileptica” (5). Epidemiological data show that the risk of dementia in PWE exceeds both that of the general population and individuals with other neurological conditions, such as stroke or migraine (6, 7). Moreover, acute symptomatic seizures following a stroke have been identified as a major risk factor for post-stroke dementia (8). Although the mechanisms underlying cognitive impairment and dementia in PWE are still unknown, emerging evidence suggests overlapping features with Alzheimer’s disease (AD) (9).

A neuropathological hallmark of AD is the aggregation of hyperphosphorylated tau (pTau) into neurofibrillary tangles (NFTs) in cell bodies and neuropil threads (NTs) in neuronal processes (10). Tau is a microtubule-associated protein that regulates microtubule stability and axonal transport (11). Accumulation of pTau can disrupt neuronal function and lead to neurodegeneration, which can clinically manifest as cognitive, behavioral, or motor symptoms (12). As abnormal phosphorylation of tau is seen in several neurodegenerative diseases, these are collectively referred to as tauopathies (11).

The presence of pTau pathology has been reported in brain specimens from PWE (13). This includes a study by Tai et al., in which accumulation of NTs and NFTs was observed in resected tissue from patients who underwent surgery for temporal lobe epilepsy (TLE). Furthermore, the pTau accumulation was found to negatively correlate with cognitive test scores at one-year post-surgery (14). Similar studies in TLE patients yielded inconsistent results (15, 16).

Thom et al. studied autopsies from patients with chronic refractory epilepsy and found a high prevalence of pTau deposits, with evidence of age-accelerated changes compared to age-matched controls. Attributing pTau accumulation to epileptic seizures was, however, difficult due to its strong association with traumatic brain injury (17), and the extent to which such comorbid tauopathies contribute to pTau pathology in PWE remains unclear. A direct link was suggested by an animal model, indicating that seizure activity itself may promote tau hyperphosphorylation in epileptogenic brain regions (18).

Inhibitory-excitatory imbalance is an important intermediate step in the early phase of AD, but evidence supporting that seizures are the dominant cause of pTau deposits remains sparse. Thus, we here studied the link between epileptic seizures and cognitive impairment in a historical cohort of patients with epilepsy and psychiatric disorders, hypothesizing that the individual brain’s pTau burden differs between the epileptogenic and non-epileptogenic hemisphere in patients with focal onset epilepsy.

### Materials and methods Patient selection

This study used human post-mortem brain tissue archived at The Danish Brain Collection located at Brain Research - Inter Disciplinary Guided Excellence (BRIDGE), University of Southern Denmark, Odense, Denmark. The brain material within this collection was obtained from autopsies conducted at Danish state mental hospitals from 1945 to 1982. We identified patients by searching for the term “epilep*” in a database of the original patient diagnoses and selected them based on information from discharge letters stored on a GDPR-secured server, both provided by the Open Patient data Explorative Network (OPEN), OUH and Department of Clinical Research, University of Southern Denmark, Odense, Denmark. Inclusion criteria were: (1) a documented history of epilepsy in accordance with the latest definition by the International League Against Epilepsy (ILAE), (2) a documented history of cognitive decline or dementia, with onset following that of epilepsy, and (3) availability of formalin-fixed brain tissue. The study was approved by the Regional Committees of Health Research Ethics for Southern Denmark (Project-ID S-20230053). The committee also granted an exemption from obtaining informed consent in accordance with section 10(1) of the Act on Research Ethics Review of Health Research Projects.

### Clinical data

Demographic and clinical data were retrieved from available medical records stored at the Danish National Archives, Copenhagen, Denmark. Recorded information regarding epilepsy included age at onset, duration, etiology, and seizure types. For patients with focal epilepsy, seizure lateralization was assessed based on seizure semiology, electroencephalographic (EEG) results, and clinical data, depending on which sources were available. For all samples, full historical pathological descriptions of the initial autopsy were available, and we noted all descriptions suggestive of vascular dementia or AD. Further, clinical details on cognitive decline and dementia were noted, too, as were any reports of traumatic head injury, stroke, psychiatric disorders, and intellectual disability.

### Immunohistochemistry

For each patient, samples were gathered from the following regions of both hemispheres: (1) the middle frontal gyrus, (2) the thalamus, and (3) the medial temporal lobe, including the hippocampal formation and parahippocampal gyrus. To ensure comparability, each sample was selected based on the availability of an anatomically similar sample from the opposite hemisphere. Immunohisto-chemistry for pTau was carried out on 3-µm thick paraffin-embedded sections mounted on charged slides. The staining was performed using the Discovery Ultra (Roche Diagnostics) automated immunostainer, following established laboratory protocols with the primary antibody anti-AT8 (1:200, Thermo Fisher Scientific, Cat# MN1020). Medial temporal lobe sections from cases without reported pathological evidence of vascular dementia, AD, or major head trauma were selected for additional amyloid-β (Aβ) staining using the anti-Aβ 4G8 antibody (1:1000, Biolegend, Cat# 800708). Sections from confirmed cases of AD were used as positive controls for the staining. All slides were digitized using a NanoZoomer slide scanner (Hamamatsu Photonics K.K.) and analyzed using NDP.view2 software (Hamamatsu Photonics K.K.). Images were acquired as snapshots from the digitized slides.

### Analysis of hyperphosphorylated tau and amyloid-β pathology

To assess the burden of pTau pathology, we created the “Tau Burden Score” (TBS). This semiquantitative scoring system was adapted from the “modified tau score” used for temporal lobe resections in Tai et al. (14). The TBS categorizes the pTau burden as none (score = 0), mild (score = 1), moderate (score = 2), or heavy (score = 3). The rater (SEL) was trained by MW and blinded to the lateralization of the likely seizure onset zone. Details of the TBS and representative images for each score are found in Supplementary Figure 1. For each section, six microscopic fields (20x) were analyzed and scored. These fields were predefined to lie within the grey matter of the sampled regions. In sections from the medial temporal lobe, the microscopic fields were distributed across the following subregions: transentorhinal cortex, entorhinal cortex, subiculum, CA1, CA2, and CA4, with one field per subregion. The adjacent tissue to each microscopic field was screened to confirm that the chosen field was representative of its area. Scores from all microscopic fields within the three sections of the same hemisphere were summed to generate a total Tau Burden Score for that hemisphere (Hemisphere-TBS). Similarly, the total Tau Burden Score of the brain (Brain-TBS) was obtained by summing the scores of both hemispheres. All scoring was performed blinded to the clinical information.

A full Braak staging was not performed, as the tissue sections did not correspond to the standard regions used for this system (19). However, if the distribution of pTau followed the Braak pattern, stages I and II were assessed. Additional presence of pTau in the middle frontal gyrus was noted as stage V, while deviations from the Braak pattern were noted as stage +. All assessed stages were evaluated according to published standards (20). Patterns of perivascular pTau aggregates or accumulation in sulcal depths were also screened for.

Aβ pathology was assessed semiquantitatively with plaque scores of none (score = 0), sparse (score = 1), moderate (score = 2), or frequent (score = 3) as outlined in the Consortium to Establish a Registry for Alzheimer’s Disease (21). Each hemisphere was analyzed and scored separately, and the higher score was used for analysis requiring a single measure per brain.

### Statistical analysis

The predefined research question was the difference in the pTau burden between the epileptogenic and non-epileptogenic hemispheres. All other comparisons were exploratory. Descriptive summaries were reported as median and interquartile range (IQR) for continuous variables, and as frequencies and percentages for categorical variables. Spearman’s (rho) correlation coefficient and the Mann-Whitney *U* test were used to examine the relationship between the pTau burden and the clinical data, as well as the Aβ plaque score. Comparison of the pTau burden and Aβ plaque score between the epileptogenic and non-epileptogenic hemispheres was assessed using the Wilcoxon signed-rank test. All tests were two-sided, and *p*-values < 0.05 were considered statistically significant. No correction for multiple testing was carried out. Statistical analyses were performed using Stata version 18 (StataCorp).

## RESULTS

### Clinical characteristics

Among 44 patients identified, 32 met the inclusion criteria and were selected for analysis. Their clinical characteristics are summarized in Table 1. The median age at death was 61.5 years (IQR = 12) and 19 patients (59.4%) were men. Epilepsy had a median duration of 39 years (IQR = 24). A known etiology was reported in six cases, including four (16%) with post-traumatic epilepsy and two (8%) with unspecified symptomatic epilepsy. A history of traumatic head injury was present in 13 patients (40.6%), seven of whom (53.8%) sustained the injury after epilepsy onset.

**Table 1.** Demographic and clinical characteristics of the patient population studied.

| Characteristics | Patients for whom data were available, *n* | Median (IQR) or *n* (%) |
| --- | --- | --- |
| Sex | 32 |  |
| Male |  | 19 (59.4) |
| Female |  | 13 (40.6) |
| Age at death, years | 32 | 61.5 (12) |
| Age at onset of epilepsy, years | 27 | 16 (20) |
| Duration of epilepsy, years | 27 | 39 (24) |
| Etiology | 25 |  |
| Cryptogenic |  | 19 (76) |
| Symptomatic |  | 2 (8) |
| Post-traumatic |  | 4 (16) |
| Seizure types | 32 |  |
| Focal onset |  | 24 (75) |
| Generalized onset |  | 8 (25) |
| Focal onset seizures | 19 |  |
| Without impaired awareness |  | 3 (15.8) |
| With impaired awareness |  | 15 (78.9) |
| Evolving to bilateral tonic-clonic seizures |  | 18 (94.7) |
| Seizure lateralization in focal epilepsy | 12 |  |
| Left |  | 7 (58.3) |
| Right |  | 5 (41.6) |
| History of traumatic brain injury | 32 | 13 (40.6) |
| History of stroke | 32 | 6 (18.6) |
| Intellectual disability | 32 | 7 (21.9) |
| History of psychiatric disorders | 32 |  |
| Psychosis |  | 19 (61.3) |
| Depression |  | 4 (6.5) |
| Behavioral dysfunction and emotional dysregulation |  | 24 (77.4) |
| Other epilepsy-related diagnoses | 32 |  |
| Psychosis epileptica |  | 6 (18.8) |
| Dementia epileptica |  | 28 (87.5) |

All patients were reported to suffer from dementia, which worsened over the course of their epilepsy. Additionally, the diagnosis of “dementia epileptica” was recorded in 28 (87.5%) cases. No further details regarding the cognitive impairment were available, except for memory loss noted in seven patients (21.9%).

### Hyperphosphorylated tau pathology

pTau pathology was observed in 22 patients (68.8%). Of these, 17 (77.3%) had both NTs and NFTs in their tissue, while 5 (22.7%) had NTs only. The median Brain-TBS was 9 (IQR = 6) within this group and 6.5 (IQR = 10.5) across all patients in the study. Figure 1A gives the Brain-TBS in the study population. Notably, two patients had a Brain-TBS that was considerably higher than the rest, with scores of 68 and 94. These two brains displayed pathological changes consistent with advanced AD at Braak stage V, as evident from both the current tissue analysis and the original histology reports (Figure 4A). Although included in the descriptive statistics, these brains were excluded from further analyses.

**Figure 1:**
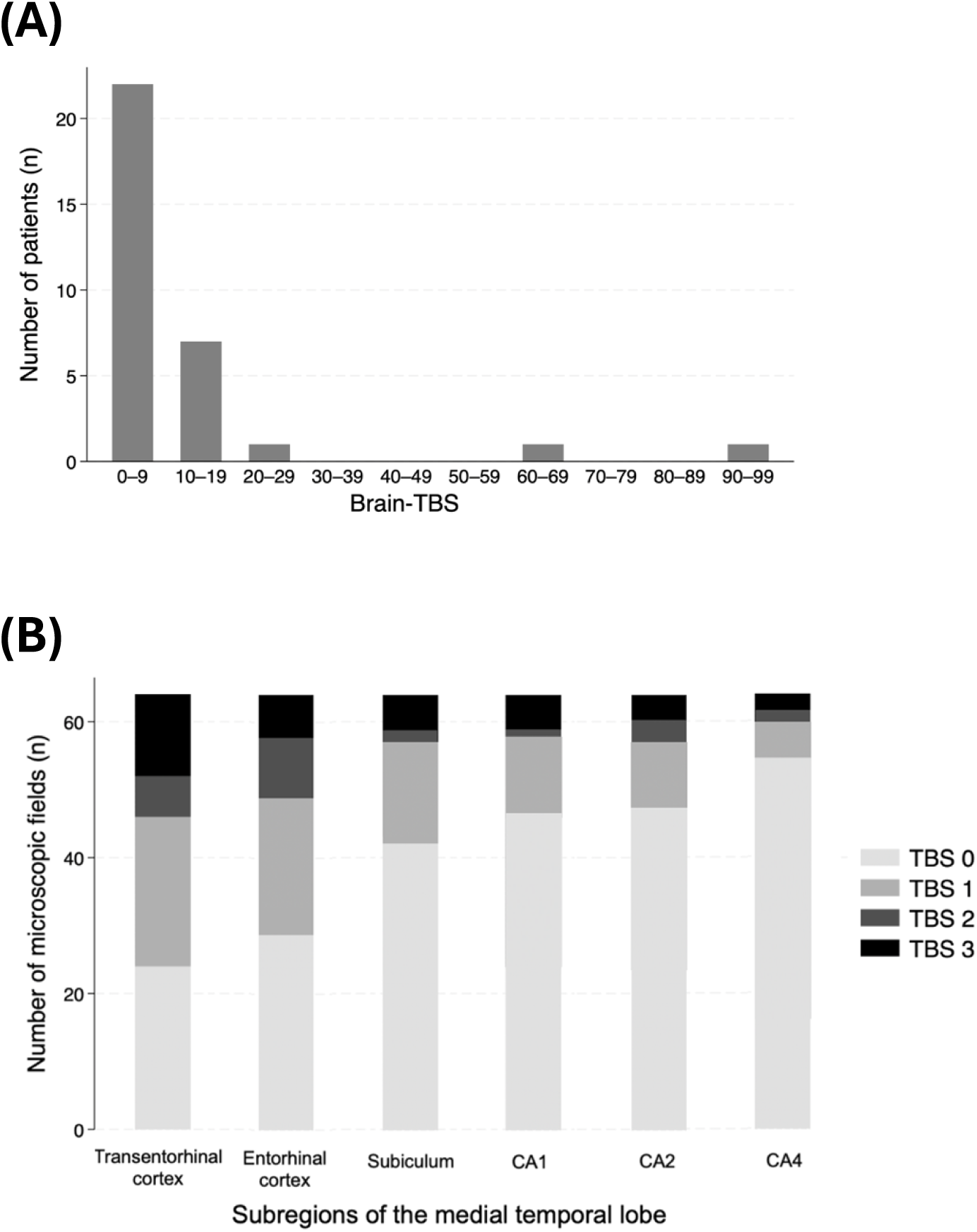
Tau burden score in the cohort. Bar charts showing the distribution of Tau Burden Scores in the study population. (**A**) The total Tau Burden Score of the brain (Brain-TBS) across all patients, where 22 out of 32 cases exhibited pTau pathology, with scores ranging from 2 to 94. (**B**) The Tau Burden Score (TBS) in microscopic fields across the subregions of the medial temporal lobe.

Sections from the medial temporal lobe were affected by pTau pathology in all 22 patients, with deposits localized exclusively to this region in 16 patients (72.3%). Figure 1B and Supplementary Table 2 gives the TBS for microscopic fields in the subregions of the medial temporal lobe. Here, the fields most frequently displayed mild pTau pathology, with 82 affected fields (59%) scoring a TBS of 1. The transentorhinal cortex was the subregion that most often displayed pTau pathology, as 39 out of 64 fields (60.9%) showed deposits, 12 of which (18.8%) were categorized as having a heavy pTau burden. In contrast, CA4 was the least affected subregion, with pTau deposits present in only nine fields (14.1%). Accumulation of pTau was identified in sections from the middle frontal gyrus and thalamus in six and three patients, respectively (27.3% and 13.6% of the 22 patients). The microscopic fields within these regions only showed mild pTau burdens when excluding the two patients with advanced AD. An overview of the TBS in each region is presented in Supplementary Table 1. Notably, pTau was bilaterally present in corresponding sections in all but one case.

Among patients displaying pathology, the distribution of pTau followed a Braak-like pattern in 11 cases (50%), including nine (40.9%) classified as stage I-II. The remaining 11 patients (50%) were classified as Braak stage +. No clear evidence of pTau accumulation around blood vessels or in sulcal depths was observed.

### Comparison of the epileptogenic and non-epileptogenic hemispheres

Seizures had a focal onset in 24 patients (75%). Available information allowed reliable lateralization in 11 brains (45.8%). Seizures most often originated from the temporal lobe, except in two patients (18.2%) with frontal lobe onset. Impaired awareness occurred in ten patients (90.9%) during seizures, including nine (81.8%) who experienced focal to bilateral tonic-clonic seizures.

In these patients, deposits of pTau were observed in the medial temporal lobe in ten brains (90.9 %), the thalamus in one brain (9.1%), and the middle frontal gyrus in one brain (9.1 %). Comparison of the hemisphere-TBS between the epileptogenic and non-epileptogenic hemispheres revealed a median score difference of 3 (IQR = 6), with a significantly higher pTau burden in the epileptogenic hemisphere (*p* = 0.02). Table 2 summarizes the seizure lateralization and hemisphere-TBS in the patients.

**Table 2.** Seizure lateralization and comparison of the Tau Burden Score in the epileptic and non-epileptic hemispheres in patients with focal onset seizures.

| Patient # | Location of the seizure onset zone | Epileptic hemisphere-TBS | Non-epileptic hemisphere-TBS | Difference in hemisphere-TBS |
| --- | --- | --- | --- | --- |
| 1 | Right temoral lobe | 1 | 2 | -1 |
| 2 | Right temporal lobe | 4 | 3 | 1 |
| 3 | Left frontal lobe | 3 | 3 | 0 |
| 4 | Right temoral lobe | 8 | 2 | 6 |
| 5 | Left temporal lobe | 47 | 47 | 0 |
| 6 | Left temoporal lobe | 6 | 2 | 4 |
| 7 | Left frontal lobe | 0 | 0 | 0 |
| 8 | Left temporal lobe | 12 | 6 | 6 |
| 9 | Right temporal lobe | 11 | 8 | 3 |
| 10 | Left temporal lobe | 14 | 6 | 8 |
| 11 | Left temporal lobe | 8 | 3 | 5 |
| 12 | Right temporal lobe | 5 | 4 | 1 |
| p-value (Wilcoxon signed-rank test) |  |  |  | 0.016 |

Abbreviations: Hemisphere-TBS; Tau Burden Score of the hemisphere.

### Associations between hyperphosphorylated tau and clinical data

The Brain-TBS correlated positively with age at death (Spearman’s rho = 0.62, *p* < 0.001) and epilepsy duration (Spearman’s rho = 0.47, *p* = 0.02, patients with AD were excluded). There was no correlation between Brain-TBS and age at epilepsy onset, and no significant associations were observed with any other patient characteristics incl. history of head trauma. Figures 2 and 3 illustrate the relationship between the Brain-TBS and the clinical variables.

**Figure 2:**
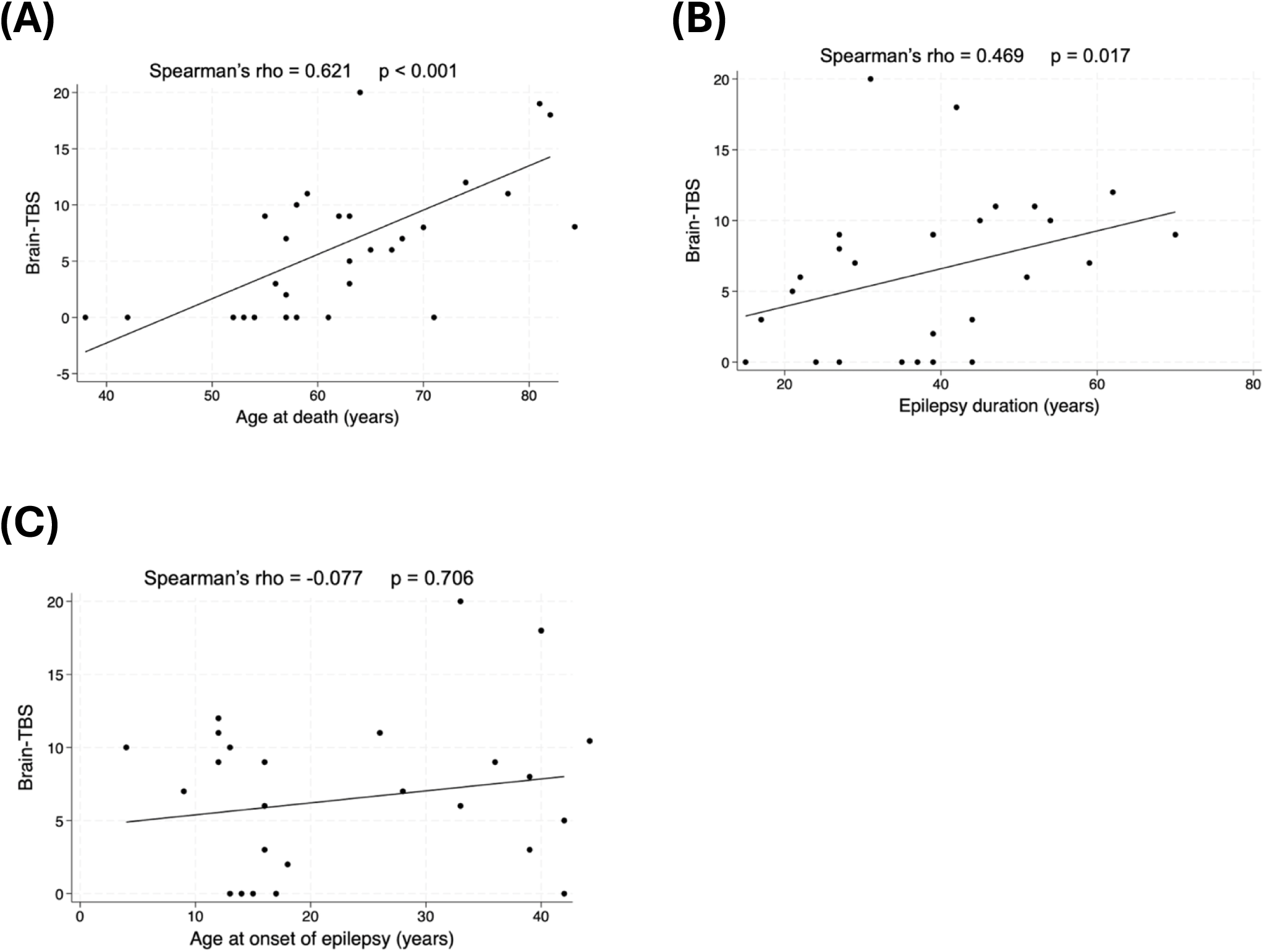
Association of the Tau burden score and clinical characteristics. (**A-C**) Association of the Tau burden score (TBS) in the brains of the patients and (**A**) the age at death, (**B**) the duration of epilepsy at the timepoint of death, and (**C**) the age of onset of epilepsy.

**Figure 3:**
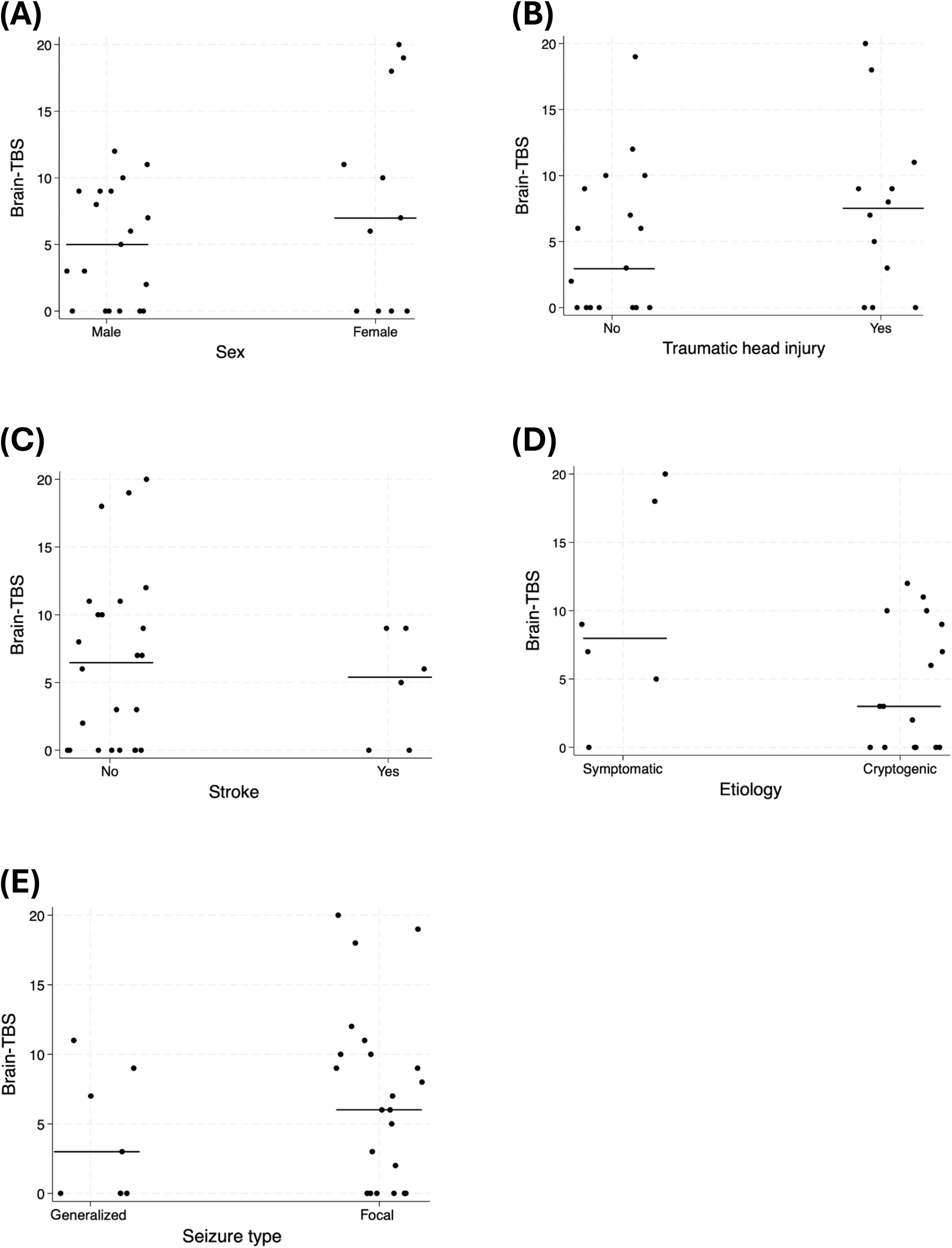
Association of the Tau burden score and clinical characteristics. Jitter plots showing the associations between the total Tau Burden Score of the brain (Brain-TBS) and categorical patient characteristics. No significant associations were observed for any of the comparisons (A-E). Horizontal lines represent the median Brain-TBS within each category.

**Figure 4:**
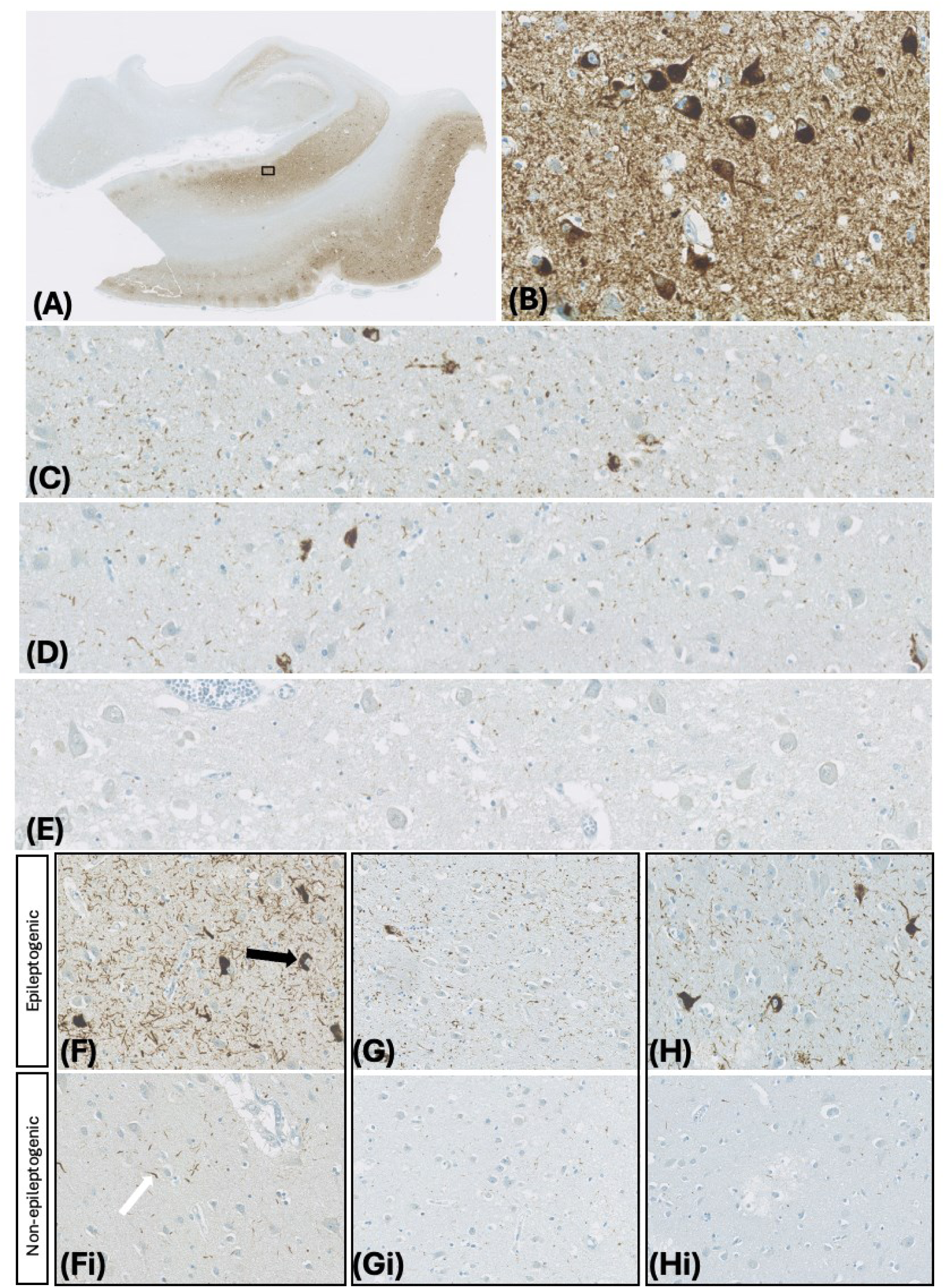
AT8 immunostaining. AT8 immunostaining of brain tissue from patients in the study, revealing hyperphosphorylated tau (pTau) pathology. pTau was observed in the form of neurofibrillary tangles (black arrowhead) and neuropil threads (white arrowhead). (**A**) Section of the medial temporal lobe from a brain with a total Tau Burden Score of 94, showing pathology consistent with Alzheimer’s disease. The rectangle is shown at higher magnification in (**B**). Overall, the burden of pTau was greater in the parahippocampal gyrus than in the hippocampus, with representative images shown from the transentorhinal cortex (**C**), entorhinal cortex (**D**), and CA1 (**E**) within a section of the medial temporal lobe with a Tau Burden Score of 7. (**F-Fi, G-Gi, H-Hi**) Examples of differences in pTau accumulation in the transentorhinal cortex between the epileptogenic and non-epileptogenic hemispheres of patients with focal onset seizures. Scale bars: (**A**) 5000 μm; (**C-E**) 250 μm; (**B, F, Fi, G, Gi, H, Hi**) 200 μm.

### Amyloid-β pathology

Eight patients showed neuropathological features suggestive of vascular dementia, including severe arteriosclerosis and multiple lacunar infarcts, while three had major brain trauma due to stroke, head injury, or tumor. The remaining nineteen patients with unknown cause of dementia were stained for Aβ. Aβ plaques were detected in eight patients (42.1%), with pathology classified as “sparse” in one brain (12.5%), “moderate” in three brains (37.5%), and “frequent” in four brains (50%), based on the highest plaque score per patient. Five Aβ-positive patients had identical plaque scores across hemispheres (62.5%), while three showed interhemispheric differences (37.5%). Supplementary Table 3 presents the Aβ plaque scores in the selected patients. In all brains with Aβ pathology, plaques were observed in the parahippocampal gyrus, whereas only three (37.5%) showed involvement of the hippocampus. No difference in Aβ plaque score was found between the epileptogenic and non-epileptogenic hemispheres (*p* = 1.00). Furthermore, no correlation was observed between Aβ plaque score and Brain-TBS (Spearman’s rho = 0.20, *p* = 0.40).

## DISCUSSION

Our findings showed aggregated pTau deposits, in the form of NTs and NFTs, in post-mortem brain specimens from patients with epilepsy who later developed dementia. While the overall extent of pTau pathology was low across cases, the pTau burden correlated significantly with both age at death and duration of epilepsy. Furthermore, a significantly higher pTau burden was observed in the epileptogenic hemisphere of patients with focal onset seizures.

Previous research has investigated the occurrence of pTau pathology in PWE, although findings have been markedly inconsistent. A recent review by Witt et al. reported that the prevalence of pTau ranged from 3.5% to 95% across five studies of patients with drug-refractory TLE (22). This wide variation may reflect differences in patient populations and in the neuropathological methodology used. The observed prevalence in the present study aligns with that reported by Thom et al., who, using the same AT8 immunostaining, identified pTau deposits in 69% of patients with characteristics similar to ours (17).

Prior studies have found no associations between pTau abundance and epilepsy-related characteristics (15, 23). In contrast, this study found the duration of epilepsy to correlate with the pTau burden, which could support the idea of seizure activity causing pTau aggregation. However, it is difficult to exclude that this correlation basically reflects an age effect rather than epilepsy itself, as most patients had early onset, life-long epilepsy. Age is a well-established risk factor for neurodegenerative diseases such as AD (7) and pTau accumulation also occurs during normal aging (24). Data from a large post-mortem series of 2,332 unselected individuals demonstrated that abnormal phosphorylated tau was present in all brains from cases aged over 50 years (25).

Nevertheless, abnormal pTau accumulation has also been reported in younger epilepsy cohorts (23, 26). Notably, Gourmaud et al. found that epilepsy patients, with a median age of 29 years, had significantly elevated pTau levels compared to age-matched controls (27). Such findings indicate that seizures might contribute to the development of pathological tau in PWE.

Traumatic brain injury has previously been identified as a strong associated factor for pTau accumulation in epilepsy, and pTau deposition patterns compatible with chronic traumatic encephalopathy (CTE) have been noted in patient tissue samples (17, 26, 28). In our study, no association was found between the pTau burden and a history of traumatic head injury. The included patients were at increased risk of seizure-related injuries, given their high frequency of seizures with impaired awareness, including bilateral tonic-clonic seizures (29). Therefore, it is possible that more cases suffered traumatic head injuries than were documented. Still, we did not observe pathological changes typical for CTE, such as perivascular or sulcal pTau foci, which argues against CTE as an underlying cause of pTau deposition our patients (30).

Most existing evidence of pTau pathology in PWE comes from surgical biopsy specimens of the temporal lobe (14, 15, 22). By using post-mortem tissue, this study was able to examine a larger portion of the brain. Nevertheless, we still found the vast majority of pTau was confined to the mesial temporal lobe. Since early-stage AD and primary age-related tauopathy are typically restricted to this region, this supports the age-related nature of the pTau deposits. On the other hand, the temporal lobe is the most frequent site of seizure onset in focal epilepsy, as was also the case in our cohort (31). Moreover, in half of the patients, the distribution of pTau deviated from the Braak-stage progression characteristic of these tauopathies (24) suggesting that seizures are contributing to pTau deposits. Unusual pTau patterns within the temporal lobe have been noted in several cases from prior epilepsy series, including reports of pTau mainly localized to the mossy fibers of the dentate gyrus (14, 28). Such findings have been interpreted as possible epilepsy-associated changes, although a distinct pTau pattern unique to epilepsy has yet to be identified.

Results from a model of intra-amygdala kainic acid-induced status epilepticus demonstrated that seizures were associated with increased pTau levels in the hippocampus ipsilateral to the injection site (32). Consistent with these findings, we observed a significantly higher pTau burden in the epileptogenic hemisphere compared to the contralateral side, supporting the concept of seizure-driven accumulation of pathological tau in the epileptic brain. The absolute asymmetry was, however, minor in the investigated subgroup, as well as the overall bilateral presence of pTau across brain sections in the study. The propagation of abnormal tau through a prion-like mechanism between neuroanatomically connected brain areas might provide an explanation (33). This is supported by data from a mesial temporal lobe epilepsy model by Canet et al. who showed that tau hyperphosphorylation occurred at the seizure focus but also extended to the contralateral hippocampus (18). Therefore, seizure-induced pTau changes may not remain strictly confined to the seizure onset zone.

In this study, we found the pTau burden to be predominantly mild across the microscopic fields, while moderate or severe pTau accumulation was rarely observed throughout all subregions within a given sample (Supplementary Table 2). Similar findings were reported by Tai et al. where mild NT staining was the most common pTau pathology detected in biopsy specimens from TLE cases (14). Likewise, Smith et al. found that only four out of 23 patients with pathological tau changes had a significant pTau burden (23). In their study, neuropsychological testing revealed moderate cognitive decline in only one patient, while the others had either normal results or mild deficits. Because cognitive impairment in AD typically emerges at Braak stage III, with dementia becoming evident at stage V-VI, the observed extent of the, possibly seizure-associated, pTau pathology alone is unlikely to account for the dementia symptoms in our cohort (34).

This could suggest that additional factors, including vascular and other brain damage as well as other protein aggregates or neuroinflammation, may contribute to cognitive decline in epilepsy. Further, the cognitive profiles reported in the patients could have been influenced by factors beyond neuropathological changes, such as anti-seizure medication, psychiatric comorbidities, or cognitive symptoms as a manifestation of seizures.

Our study has limitations. Although immunostaining is a well-established method for identifying pTau pathology, the analysis typically relies on observer-dependent visual assessment. Furthermore, we created a new scoring system, which has not been formally validated, although it resembles the semiquantitative approaches used in similar studies. We acknowledge that using tissue samples stored in formalin for several decades may influence pTau immunohistochemistry due to loss of antigenicity or degradation of the proteins, which could have contributed to the low or absent pTau presence in some cases. Another challenge from working with a historical tissue collection was the incomplete clinical data, as several patients lacked detailed documentation of their medical histories. Inherent in most studies in this field, we were also limited by the relatively small number of patients available.

In conclusion, the “dementia epileptica” diagnosis in this cohort was associated with increased pTau burden in the epileptogenic hemisphere and time since epilepsy diagnosis, supporting the concept of seizure-induced pTau accumulation. However, pTau is unlikely to be the primary neuropathological link between epilepsy and dementia in the cohort studied, given the overall mild pathology and the presence of competing diagnoses.

## ACHNOWLEDGEMENTS

The authors are grateful to Lone Christiansen for her help in performing the immunostaining and the Danish National Archives for providing access to the patients’ charts.

## COMPETING INTERESTS

None of the authors had any conflict of interest to disclose relevant to this study. We confirm that we have read the Journal’s position on issues involved in ethical publication and affirm that this report is consistent with those guidelines.

## AVAILABILITY OF DATA AND MATERIALS

All data generated or analysed during this study are included in this published article and its supplementary information files.

## FUNDING

This study was supported by The Lundbeck Foundation (grant ID: 6214271 and R434-2023-342) and the University Hospital Odense (BRIDGE collaboration).

## AUTHOR CONTRIBUTIONS

SEL analyzed all histology, wrote the draft of the manuscript and interpreted patient data supported by CDC. JP supervised and facilitated the histological staining performed as part of this study. MW contributed to the design of the study, selection of patients, and quality assurance of the staining. CPB designed and supervised the study, secured funding, and finalized the draft of the manuscript. All authors read and approved the final manuscript.

## Data Availability

All data generated or analysed during this study are included in this published article and its supple-mentary information files.

## Notes

### Competing Interest Statement

The authors have declared no competing interest.

### Author Declarations

The study was approved by the Regional Committees of Health Research Ethics for Southern Den-mark (Project-ID S-20230053). The committee also granted an exemption from obtaining informed consent in accordance with section 10(1) of the Act on Research Ethics Review of Health Research Projects.

## REFERENCES

1. WHO. Epilepsy: a public health imperative. Geneva; 2019.

2. Thijs RD, Surges R, O’Brien TJ, Sander JW. Epilepsy in adults. Lancet. 2019;393(10172):689–701.

3. Keezer MR, Sisodiya SM, Sander JW. Comorbidities of epilepsy: current concepts and future perspectives. Lancet Neurol. 2016;15(1):106–15.

4. Hoxhaj P, Habiya SK, Sayabugari R, Balaji R, Xavier R, Ahmad A, et al. Investigating the Impact of Epilepsy on Cognitive Function: A Narrative Review. Cureus. 2023;15(6):e41223.

5. Helmstaedter C, Witt JA. Epilepsy and cognition - A bidirectional relationship? Seizure. 2017;49:83–9.

6. Tai XY, Torzillo E, Lyall DM, Manohar S, Husain M, Sen A. Association of Dementia Risk With Focal Epilepsy and Modifiable Cardiovascular Risk Factors. JAMA Neurol. 2023;80(5):445–54.

7. Zhao N, Chen H, Zhang W, Yao J, Tu Q, Yu X, Sun X. Bidirectional influences between seizures and dementia: A systematic review and meta-analysis. Int J Geriatr Psychiatry. 2022;37(7).

8. Lekoubou A, Ba DM, Nguyen C, Liu G, Leslie DL, Bonilha L, Vernon CM. Poststroke Seizures and the Risk of Dementia Among Young Stroke Survivors. Neurology. 2022;99(4):e385–e92.

9. Lu O, Kouser T, Skylar-Scott IA. Alzheimer’s disease and epilepsy: shared neuropathology guides current and future treatment strategies. Front Neurol. 2023;14:1241339.

10. Braak H, Braak E. Neuropathological stageing of Alzheimer-related changes. Acta Neuropathol. 1991;82(4):239–59.

11. Creekmore BC, Watanabe R, Lee EB. Neurodegenerative Disease Tauopathies. Annu Rev Pathol. 2024;19:345–70.

12. Zhang Y, Wu KM, Yang L, Dong Q, Yu JT. Tauopathies: new perspectives and challenges. Mol Neurodegener. 2022;17(1):28.

13. Sánchez MP, García-Cabrero AM, Sánchez-Elexpuru G, Burgos DF, Serratosa JM. Tau-Induced Pathology in Epilepsy and Dementia: Notions from Patients and Animal Models. Int J Mol Sci. 2018;19(4).

14. Tai XY, Koepp M, Duncan JS, Fox N, Thompson P, Baxendale S, et al. Hyperphosphorylated tau in patients with refractory epilepsy correlates with cognitive decline: a study of temporal lobe resections. Brain. 2016;139(Pt 9):2441–55.

15. Aroor A, Nguyen P, Li Y, Das R, Lugo JN, Brewster AL. Assessment of tau phosphorylation and β-amyloid pathology in human drug-resistant epilepsy. Epilepsia Open. 2023;8(2):609–22.

16. Silva JC, Vivash L, Malpas CB, Hao Y, McLean C, Chen Z, et al. Low prevalence of amyloid and tau pathology in drug-resistant temporal lobe epilepsy. Epilepsia. 2021;62(12):3058–67.

17. Thom M, Liu JY, Thompson P, Phadke R, Narkiewicz M, Martinian L, et al. Neurofibrillary tangle pathology and Braak staging in chronic epilepsy in relation to traumatic brain injury and hippocampal sclerosis: a post-mortem study. Brain. 2011;134(Pt 10):2969–81.

18. Canet G, Zub E, Zussy C, Hernandez C, Blaquiere M, Garcia V, et al. Seizure activity triggers tau hyperphosphorylation and amyloidogenic pathways. Epilepsia. 2022;63(4):919–35.

19. Braak H, Alafuzoff I, Arzberger T, Kretzschmar H, Del Tredici K. Staging of Alzheimer disease-associated neurofibrillary pathology using paraffin sections and immunocytochemistry. Acta Neuropathol. 2006;112(4):389–404.

20. Alafuzoff I, Arzberger T, Al-Sarraj S, Bodi I, Bogdanovic N, Braak H, et al. Staging of neurofibrillary pathology in Alzheimer’s disease: a study of the BrainNet Europe Consortium. Brain Pathol. 2008;18(4):484–96.

21. Mirra SS, Heyman A, McKeel D, Sumi SM, Crain BJ, Brownlee LM, et al. The Consortium to Establish a Registry for Alzheimer’s Disease (CERAD). Part II. Standardization of the neuropathologic assessment of Alzheimer’s disease. Neurology. 1991;41(4):479–86.

22. Witt JA, Andernach J, Becker A, Helmstaedter C. Hyperphosphorylated Tau and Cognition in Epilepsy. J Clin Med. 2025;14(2).

23. Smith KM, Blessing MM, Parisi JE, Britton JW, Mandrekar J, Cascino GD. Tau deposition in young adults with drug-resistant focal epilepsy. Epilepsia. 2019;60(12):2398–403.

24. Crary JF, Trojanowski JQ, Schneider JA, Abisambra JF, Abner EL, Alafuzoff I, et al. Primary age-related tauopathy (PART): a common pathology associated with human aging. Acta Neuropathol. 2014;128(6):755–66.

25. Braak H, Thal DR, Ghebremedhin E, Del Tredici K. Stages of the pathologic process in Alzheimer disease: age categories from 1 to 100 years. J Neuropathol Exp Neurol. 2011;70(11):960–9.

26. Puvenna V, Engeler M, Banjara M, Brennan C, Schreiber P, Dadas A, et al. Is phosphorylated tau unique to chronic traumatic encephalopathy? Phosphorylated tau in epileptic brain and chronic traumatic encephalopathy. Brain Res. 2016;1630:225–40.

27. Gourmaud S, Shou H, Irwin DJ, Sansalone K, Jacobs LM, Lucas TH, et al. Alzheimer-like amyloid and tau alterations associated with cognitive deficit in temporal lobe epilepsy. Brain. 2020;143(1):191–209.

28. Ryniejska M, El-Hachami H, Mrzyglod A, Liu J, Thom M. The prevalence of chronic traumatic encephalopathy in a historical epilepsy post-mortem collection. Brain Pathol. 2025;35(3):e13317.

29. Asadi-Pooya AA, Nikseresht A, Yaghoubi E, Nei M. Physical injuries in patients with epilepsy and their associated risk factors. Seizure. 2012;21(3):165–8.

30. Bieniek KF, Cairns NJ, Crary JF, Dickson DW, Folkerth RD, Keene CD, et al. The Second NINDS/NIBIB Consensus Meeting to Define Neuropathological Criteria for the Diagnosis of Chronic Traumatic Encephalopathy. J Neuropathol Exp Neurol. 2021;80(3):210–9.

31. Chowdhury FA, Silva R, Whatley B, Walker MC. Localisation in focal epilepsy: a practical guide. Pract Neurol. 2021;21(6):481–91.

32. Alves M, Kenny A, de Leo G, Beamer EH, Engel T. Tau Phosphorylation in a Mouse Model of Temporal Lobe Epilepsy. Front Aging Neurosci. 2019;11:308.

33. Mudher A, Colin M, Dujardin S, Medina M, Dewachter I, Alavi Naini SM, et al. What is the evidence that tau pathology spreads through prion-like propagation? Acta Neuropathol Commun. 2017;5(1):99.

34. Therriault J, Pascoal TA, Lussier FZ, Tissot C, Chamoun M, Bezgin G, et al. Biomarker modeling of Alzheimer’s disease using PET-based Braak staging. Nat Aging. 2022;2(6):526–35.

