## Supplementary Table for "The neuropathology of “dementia epileptica”"

**Supplementary Figure 1. Visual representation of the Tau Burden Score**

TBS 0

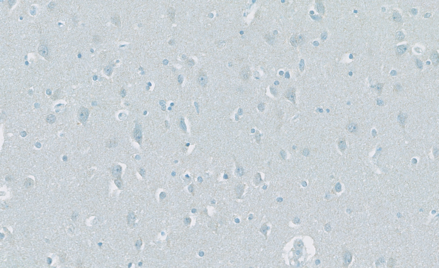

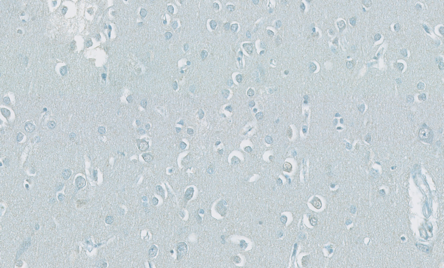

No AT8-labeled NTs or NFTs

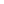

TBS 1

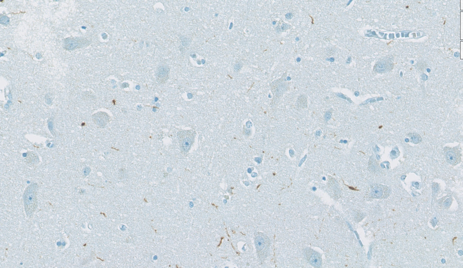

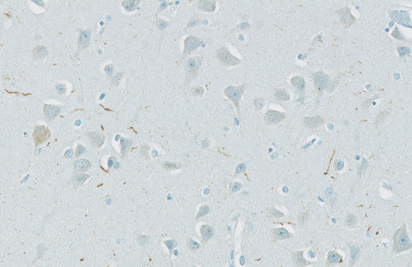

Mild burden of AT8-labeled NTs at 20x magnification

TBS 2

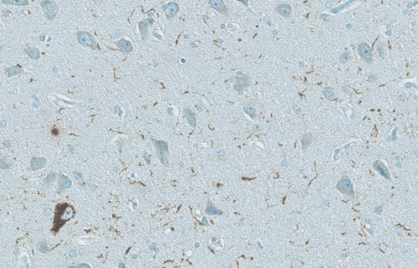

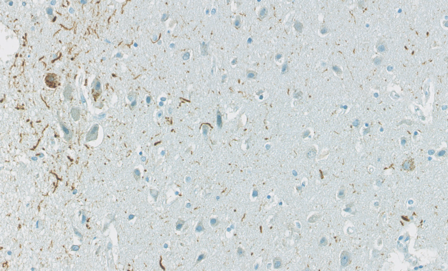

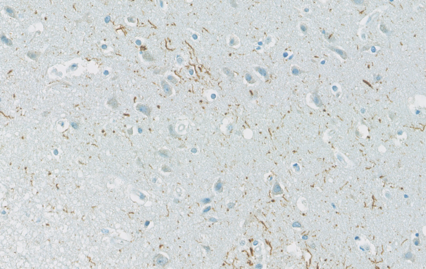

Sporadic AT8-labeled NFTs at 20x magnification (1-2 NFTs within the microscopic field)

Moderate burden of AT8-labeled NTs at 20x magnification

and/or

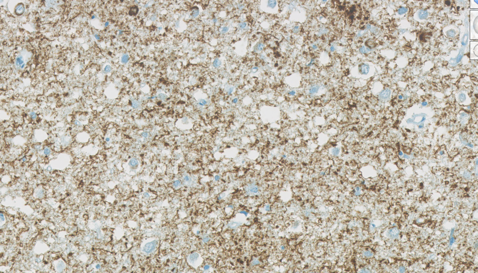

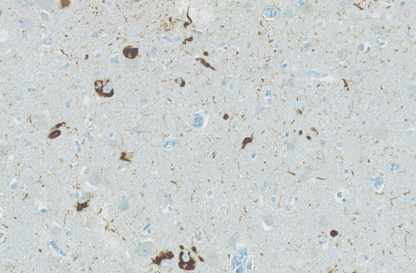

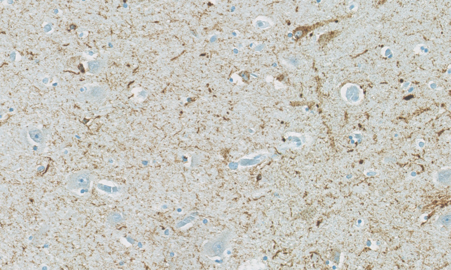
TBS 3

Accumulation of AT8-labeled NFTs at 20x magnification

Heavy burden of AT8-labeled NTs at 20x magnification

and/or

Abbreviations: TBS, Tau Burden Score; NTs, neuropil threads; NFTs, neurofibrillary tangles

**Supplementary Table 1**. **Tau Burden Score description**

| **Tau Burden Score** | **Description of NTs** |  | **Description of NFTs** |
| --- | --- | --- | --- |
| 0 | No AT8 labeled NTs |  | No AT8-labeled NFTs |
| 1 | Mild burden of AT8-labeled NTs at 20x magnification (barely present at 10x magnification) |  | No AT8-labeled NFTs |
| 2 | Moderate burden of AT8-labeled NTs at 20x magnification (easily noted at 10x magnification) | and/or | Sporadic AT8-labeled NFTs at 20x magnification (1-2 NFTs within the microscopic field) |
| 3 | Heavy burden of AT8-labeled NTs at 20x magnification (can be visualized without any magnification) | and/or | Accumulation of AT8-labeled NFTs at 20x magnification |

Abbreviations: NTs, neuropil threads; NFTs, neurofibrillary tangles

| **Patient #** | **TBS left MTL** | **TBS left thalamus** | **TBS left MFG** | **Hemisphere-TBS left** | **TBS right MTL** | **TBS right thalamus** | **TBS right MFG** | **Hemisphere-TBS right** | **Brain-TBS** |
| --- | --- | --- | --- | --- | --- | --- | --- | --- | --- |
| **1** | 2 | 0 | 0 | 2 | 1 | 0 | 0 | 1 | 3 |
| **2** | 3 | 0 | 0 | 3 | 4 | 0 | 0 | 4 | 7 |
| **3** | 3 | 0 | 0 | 3 | 3 | 0 | 0 | 3 | 6 |
| **4** | 2 | 0 | 0 | 2 | 8 | 0 | 0 | 8 | 10 |
| **5** | 6 | 0 | 0 | 6 | 2 | 0 | 0 | 2 | 8 |
| **6** | 0 | 0 | 0 | 0 | 0 | 0 | 0 | 0 | 0 |
| **7** | 9 | 3 | 0 | 12 | 4 | 2 | 0 | 6 | 18 |
| **8** | 7 | 0 | 1 | 8 | 10 | 0 | 1 | 11 | 19 |
| **9** | 14 | 0 | 0 | 14 | 6 | 0 | 0 | 6 | 20 |
| **10** | 8 | 0 | 0 | 8 | 3 | 0 | 0 | 3 | 11 |
| **11** | 4 | 0 | 0 | 4 | 5 | 0 | 0 | 5 | 9 |
| **12** | 17 | 12 | 18 | 47 | 17 | 12 | 18 | 47 | 94 |
| **13** | 6 | 0 | 0 | 6 | 1 | 0 | 0 | 1 | 7 |
| **14** | 4 | 0 | 0 | 4 | 2 | 0 | 0 | 2 | 6 |
| **15** | 10 | 0 | 0 | 10 | 2 | 0 | 0 | 2 | 12 |
| **16** | 7 | 0 | 0 | 7 | 4 | 0 | 0 | 4 | 11 |
| **17** | 2 | 0 | 0 | 2 | 0 | 0 | 0 | 0 | 2 |
| **18** | 1 | 0 | 0 | 1 | 2 | 0 | 0 | 2 | 3 |
| **19** | 3 | 0 | 4 | 7 | 2 | 0 | 1 | 3 | 10 |
| **20** | 3 | 0 | 2 | 5 | 2 | 0 | 2 | 4 | 9 |
| **21** | 1 | 0 | 1 | 2 | 2 | 0 | 5 | 7 | 9 |
| **22** | 18 | 4 | 8 | 30 | 18 | 5 | 15 | 38 | 68 |
| **23** | 2 | 0 | 0 | 2 | 3 | 0 | 0 | 3 | 5 |
| **24** | 0 | 0 | 0 | 0 | 0 | 0 | 0 | 0 | 0 |
| **25** | 0 | 0 | 0 | 0 | 0 | 0 | 0 | 0 | 0 |
| **26** | 0 | 0 | 0 | 0 | 0 | 0 | 0 | 0 | 0 |
| **27** | 0 | 0 | 0 | 0 | 0 | 0 | 0 | 0 | 0 |
| **28** | 0 | 0 | 0 | 0 | 0 | 0 | 0 | 0 | 0 |
| **29** | 0 | 0 | 0 | 0 | 0 | 0 | 0 | 0 | 0 |
| **30** | 0 | 0 | 0 | 0 | 0 | 0 | 0 | 0 | 0 |
| **31** | 0 | 0 | 0 | 0 | 0 | 0 | 0 | 0 | 0 |
| **32** | 0 | 0 | 0 | 0 | 0 | 0 | 0 | 0 | 0 |

Abbreviations: TBS, Tau Burden Score; MTL, medial temporal lobe; MGF, middle frontal gyrus

**Supplementary Table 2. Overview of the Tau Burden Score of each brain region, hemisphere, and whole brain in the study population**

| **Patient #** | **Left TEC** | **Left EC** | **Left Sub** | **Left CA1** | **Left CA2** | **Left CA4** | **Right TEC** | **Right EC** | **Right Sub** | **Right CA1** | **Right CA2** | **Right CA4** |
| --- | --- | --- | --- | --- | --- | --- | --- | --- | --- | --- | --- | --- |
| **1** | 1 | 1 | 0 | 0 | 0 | 0 | 1 | 0 | 0 | 0 | 0 | 0 |
| **2** | 1 | 2 | 0 | 0 | 0 | 0 | 2 | 2 | 0 | 0 | 0 | 0 |
| **3** | 1 | 1 | 0 | 1 | 0 | 0 | 1 | 1 | 1 | 0 | 0 | 0 |
| **4** | 1 | 1 | 0 | 0 | 0 | 0 | 3 | 3 | 1 | 1 | 0 | 0 |
| **5** | 2 | 1 | 1 | 1 | 1 | 0 | 1 | 1 | 0 | 0 | 0 | 0 |
| **6** | 0 | 0 | 0 | 0 | 0 | 0 | 0 | 0 | 0 | 0 | 0 | 0 |
| **7** | 3 | 2 | 1 | 1 | 1 | 1 | 1 | 1 | 1 | 0 | 1 | 0 |
| **8** | 1 | 1 | 1 | 1 | 1 | 1 | 2 | 2 | 2 | 1 | 2 | 1 |
| **9** | 3 | 2 | 3 | 3 | 2 | 1 | 1 | 1 | 1 | 1 | 1 | 1 |
| **10** | 3 | 3 | 1 | 1 | 0 | 0 | 1 | 1 | 1 | 0 | 0 | 0 |
| **11** | 3 | 1 | 0 | 0 | 0 | 0 | 3 | 1 | 1 | 0 | 0 | 0 |
| **12** | 3 | 3 | 3 | 3 | 2 | 3 | 3 | 3 | 3 | 3 | 3 | 2 |
| **13** | 3 | 2 | 0 | 1 | 0 | 0 | 1 | 0 | 0 | 0 | 0 | 0 |
| **14** | 2 | 2 | 0 | 0 | 0 | 0 | 1 | 1 | 0 | 0 | 0 | 0 |
| **15** | 2 | 2 | 2 | 2 | 2 | 0 | 0 | 1 | 0 | 0 | 1 | 0 |
| **16** | 3 | 2 | 1 | 1 | 0 | 0 | 1 | 0 | 1 | 1 | 1 | 0 |
| **17** | 1 | 1 | 0 | 0 | 0 | 0 | 0 | 0 | 0 | 0 | 0 | 0 |
| **18** | 0 | 1 | 0 | 0 | 0 | 0 | 0 | 1 | 1 | 0 | 0 | 0 |
| **19** | 2 | 1 | 0 | 0 | 0 | 0 | 0 | 0 | 1 | 0 | 1 | 0 |
| **20** | 1 | 1 | 0 | 0 | 1 | 0 | 1 | 1 | 0 | 0 | 0 | 0 |
| **21** | 1 | 0 | 0 | 0 | 0 | 0 | 1 | 1 | 0 | 0 | 0 | 0 |
| **22** | 3 | 3 | 3 | 3 | 3 | 3 | 3 | 3 | 3 | 3 | 3 | 3 |
| **23** | 1 | 0 | 1 | 0 | 0 | 0 | 1 | 0 | 1 | 1 | 0 | 0 |
| **24** | 0 | 0 | 0 | 0 | 0 | 0 | 0 | 0 | 0 | 0 | 0 | 0 |
| **25** | 0 | 0 | 0 | 0 | 0 | 0 | 0 | 0 | 0 | 0 | 0 | 0 |
| **26** | 0 | 0 | 0 | 0 | 0 | 0 | 0 | 0 | 0 | 0 | 0 | 0 |
| **27** | 0 | 0 | 0 | 0 | 0 | 0 | 0 | 0 | 0 | 0 | 0 | 0 |
| **28** | 0 | 0 | 0 | 0 | 0 | 0 | 0 | 0 | 0 | 0 | 0 | 0 |
| **29** | 0 | 0 | 0 | 0 | 0 | 0 | 0 | 0 | 0 | 0 | 0 | 0 |
| **30** | 0 | 0 | 0 | 0 | 0 | 0 | 0 | 0 | 0 | 0 | 0 | 0 |
| **31** | 0 | 0 | 0 | 0 | 0 | 0 | 0 | 0 | 0 | 0 | 0 | 0 |
| **32** | 0 | 0 | 0 | 0 | 0 | 0 | 0 | 0 | 0 | 0 | 0 | 0 |

Abbreviations: TEC, transentorhinal cortex; EC, entorhinal cortex; Sub, subiculum.

**Supplementary Table 3. Overview of the Tau Burden Score of microscopic fields in the subregions of the medial temporal lobe**

**Supplementary Table 3. Overview of the amyloid-β plaque score of each hemisphere**

| **Patient #** | **Left hemisphere** | **Right hemisphere** |
| --- | --- | --- |
| **3** | 0 | 0 |
| **4** | 2 | 3 |
| **5** | 1 | 3 |
| **6** | 2 | 2 |
| **7** | 0 | 0 |
| **8** | 2 | 2 |
| **9** | 0 | 0 |
| **10** | 3 | 3 |
| **13** | 0 | 0 |
| **15** | 0 | 0 |
| **16** | 3 | 3 |
| **17** | 0 | 0 |
| **18** | 2 | 1 |
| **19** | 0 | 0 |
| **24** | 0 | 0 |
| **25** | 0 | 0 |
| **26** | 1 | 1 |
| **27** | 0 | 0 |
| **28** | 0 | 0 |
